# Face-masking, an acceptable protective measure against COVID-19 - Findings of Ugandan high-risk groups

**DOI:** 10.1101/2020.08.29.20184325

**Authors:** Gerald Mboowa, David Musoke, Douglas Bulafu, Dickson Aruhomukama

## Abstract

Face-masking could reduce the risk of COVID-19 transmission. We assessed: knowledge, attitudes, perceptions, and practices towards COVID-19 and face-mask use among 644 high risk-individuals in Kampala, Uganda. In data analysis, descriptive, bivariate and multivariate logistic regression analyses, with a 95% confidence interval were considered. Adjusted-odds ratios were used to determine the magnitude of associations. P-values < 0.05 were considered statistically-significant. Majority: 99.7% and 87.3% of the participants respectively had heard and believed that face-masks were protective against COVID-19, while 67.9% reported having received information on face-mask use. Males, food market vendors, those with no formal education, and those aged 24-33, 44-53 and 54-63 years were 0.58, 0.47, 0.25, 1.9, 2.12, and 3.39 times less likely to have received information about face-mask use respectively. Majority, 67.8% owned locally-made, non-medical face-masks, while 77.0% of face-mask owners believed that they knew the right procedure of wearing them. Those who had received information on face-mask use were 2.85 and 1.83 times more likely to own face-masks and to perceive them as protective. Food market vendors were 3.92 times more likely to re-use their face-masks. Our findings suggest that Ugandan high-risk groups have good knowledge, optimistic attitudes and perceptions, and relatively appropriate practices towards COVID-19.

## Introduction

Coronavirus disease 2019 (COVID-19) is an acute-respiratory infectious disease caused by the Severe Acute Respiratory Syndrome Coronavirus 2 (SARS-CoV-2), that spreads mainly through respiratory droplets and secretions (1,2). The disease was first reported in Wuhan, Hubei Province of China in December 2019 (3). COVID-19 transmission can occur directly via contact with infected individuals or indirectly via contact with surfaces in their immediate environment or objects used on or by those infected (1,4—6). In specific circumstances and settings particularly where procedures that generate aerosols are performed, airborne transmission of COVID-19 could be possible (7-9). The spread of COVID-19 via aerosols even in the absence of aerosol generating procedures could also be possible (7-9). To date, no clear treatment options have been reported for the virus and as such, treatments have been limited to the use of anti-HIV drugs and/or other anti-virals such as Galidesivir and Remdesivir (10).

To contain viral spread, several countries continue to utilize non-pharmaceutical public health interventions (11-13), including among others: (1) boarder control or closure, (2) partial- or complete-lockdown, (3) quarantine and testing of incoming travelers and returnees, and (4) mass testing for rapid case detection, contact tracing and quarantine (10). Additional measures, community mitigation strategies including among others: (1) mass media-based sensitization, (2) appealing to the masses to: unceasingly carry out good hygiene practices particularly hand washing, maintain appropriate social distance, stop all mass gatherings, cease all socioeconomic activities except essential services like security, food markets, health-care and wear face-masks also continue to be emphasized (10,14,15).

These measures have been implemented at different time points and to various degrees in different geographical areas to reduce the risk of community transmission of COVID-19 (10,14,15). Noteworthy, several of these measures had been used previously for the control of community transmission of the: (1) Severe Acute Respiratory Syndrome (SARS) in 2003, (2) pandemic Influenza A H1N1 in 2009 (2,16,17), Ebola Viral Hemorrhagic Fever in West Africa in 2014 (18,19), as well as several viral hemorrhagic fever outbreaks over the years in Uganda (20).

Wearing of face-masks in public settings where social distancing measures are difficult to maintain, has been documented as one of the most critical prevention measure that can limit the acquisition and spread of COVID-19 by the World Health Organization (WHO) and the United States Centers for Disease Control (CDC). In light of this, WHO and CDC have developed guidelines for the use of the same in these settings (21,22).

Previously published studies have shown that wearing of face-masks to control infectious diseases spread has several advantages that include among others: (1) simple operation, (2) strong sustainability, (3) high health benefits, and (4) good health economic benefits (23-25). Other previously published studies have also shown that use of face-masks by the general public is of potentially high value in limiting community transmission of infectious diseases (2,26-28). Likewise, the use of face-masks has also been documented to curb viral transmission by asymptomatic individuals and thus limiting the epidemic’s growth rate (28). With regards to limiting community spread of COVID-19, community-wide use of face-masks has been encouraged (29,30). Face-masks have also been suggested to serve as visible cues of an otherwise yet widely prevalent pathogen, SARS-CoV-2, and as tools that could be utilized to remind people of the importance of the other infection-control measures such as social distancing (31). Face-masks have also been documented to be symbolic, beyond them being tools, they have been described as talismans that could increase health-care workers’ perceived sense of safety, well-being, and trust in their health-care settings (31). Similar to a few other countries, Uganda continues to implement a phased-approach of lifting the countrywide lockdown while considering the wearing of face-masks in all public settings mandatory for all (32). In light of this, we hypothesized that high knowledge levels about COVID-19 and face-mask use, positive attitudes and perceptions towards face-mask use as well as good face-mask use practices in Uganda could significantly contribute to breaking the chain of SARS-CoV-2 transmission in health-care settings and the community via reducing the infectiousness of the sub-clinical virus shedders while also offering some protection to the susceptible populations. Hence, we aimed to provide evidence on health-care and community-level perspectives on the use of face-masks in preventing COVID-19 acquisition and spread through assessing the knowledge, attitudes, perceptions, and practices towards their use. This is because little remains known regarding use of face-masks in Uganda. We hoped that our findings would be used by decision makers to guide their recommendations with regards to the use of face-masks by the population including healthy, pre-symptomatic and asymptomatic individuals in health-care settings and the community to prevent health-care settings and community acquisition and spread of COVID-19.

## Materials and Methods

### Study design

This study was a cross-sectional study, and was part of a larger study titled: Assessing knowledge, attitudes, perceptions and skills towards the use of face-masks: a community-level perspective (MASKUG-2020), that is aimed to assess: (1) knowledge, attitudes, perceptions, practices, and skills towards the use of face-masks by high-risk groups in Kampala district, Uganda, (2) skills towards the use of face-masks by the same groups, and (3) to evaluate the face-masks for safety and fitness-for-use, (4) to provide a classification for those commonly circulating on the Ugandan market, as well as (5) to educate and skill the same groups on the rational use and disposal of face-masks.

### Study sites and settings

This study was carried out in Kampala, the capital city of Uganda. Kampala is divided into five divisions namely: Central, Kawempe, Makindye, Nakawa, and Rubaga. Study sites were purposively selected to represent these five divisions and included: (1) food markets namely: (i) Owino market located in downtown Kampala, (ii) Nakasero market located at the foot of Nakasero hill, (iii) Bugolobi market located along Old-Portbell road, (iv) Nakawa market located along Kampala-Jinja highway, (v) Kalerwe market located on the Kampala Northern by-pass along Gayaza road, (vii) Kasubi market located along Kampala-Hoima road, and (viii) Wandegeya market located in front of the four-way junction North and North East of Makerere University, East and North of Mulago National Referral Hospital, and South and South East of Nakasero hill (33), (2) police stations namely: Central, Old-Kampala, Katwe, Mulago, Kanyanya and Wandegeya, and (3) Mulago National Referral Hospital, the largest public hospital in Uganda located on Mulago hill in the Northern part of the city Kampala.

### Study population and period

The study population comprised high-risk groups namely: (1) food market vendors that included food store owners and sellers of fruits and vegetables, (2) police officers mainly traffic officers and curfew enforcers, (3) health-care workers mainly nurses and medical doctors. All these had been allowed to continue their businesses during the entire countrywide lockdown ordered by the Ugandan government; this was because they were considered as essential service providers. The study was conducted in July 2020. High-risk groups in this study constituted persons who were working in crowded sites, who continuously interacted with multiple different people on a day-to-day basis which meant that they were at a higher risk of contracting and/or transmitting COVID-19, as urban crowding has been documented to influence the overall load of infectious agents, including respiratory viruses (34,35).

### Sample size and sampling

This study’s sample size constituted 659 study participants. The sample size was calculated using the Kish Leslie formula (1995) for cross-sectional studies giving a sample size of 384. Since most of the targeted participants were working in shifts, we considered a non-response rate of 30%, and a design effect of 1.2 (36), giving us a sample size of 659 study participants. At each of the sites, multi-stage sampling was done based on the average number of participants present to ensure equal representation of all the sites. Several clusters of 20-25 participants were selected from each site using the probability proportion to size sampling, which ensured that all individuals in the target populations had an equal chance of being selected. Three to four busy days of the week were purposively selected to visit each of the sites.

### Questionnaire design

A semi-structured questionnaire based on the Occupational Safety and Health Administration (OSHA) Respiratory Protection Program standard requirements (OSHA, 2017) and the Ministry of Health in partnership with UNICEF-KAP survey on COVID-19-Response (37)s, was developed and used in the data collection. One occupational/environmental health and safety expert, a statistician, and three health-care workers (i.e. one doctor and two nurses) assessed the validity of the questionnaire. The reliability of the questionnaire was checked by Cronbach’s alpha *(α =* 0.860, 0.899 and 0.870, respectively, for knowledge, attitudes and perceptions, and practices dimensions). The questionnaire consisted of five components including: demographics, knowledge (12 items), attitudes, perceptions, and practices (10 items). Knowledge items were categorized as yes (score 1) and no (score 0). Attitudes, perceptions, and practices items were scored using a Likert-scales, which ranged from 1 (very fearful) to 4 (optimistic) and 1 (strongly agree) to 4 (strongly disagree). Other attitudes, perceptions, and practices items were categorized as yes (score 1) and no (score 0). All negatively worded responses were scored reversely. In addition, the study questionnaire was evaluated for face and internal validity by the principal investigators. Also, to enhance data quality, all research assistants (RAs) were trained and supervised, and the questionnaire was pre-tested.

### Data collection, validation and analysis

Data was collected by the trained RAs using the developed semi-structured questionnaires. Briefly, the data was entered using mobile android and iOS phones and tablets. These had been loaded with the Open Data Kit application (ODK), and the data was synchronized onto a remote server daily. Data collection using mobile android and iOS phones and tablets allowed for real-time data capture and entry, minimized errors at entry and eased data cleaning. To ensure that the data was secure, only the principal investigators had the security key to the ODK server, where the data was being sent during data collection. Validation of the collected data was done by checking a significant percentage (20 - 30%) of the same by field supervisors and the principal investigators. The collected data was cleaned using MS Excel 2016 and analyzed using STATA 14.0 statistical software. Descriptive analyses such as frequencies, proportions, and means (where appropriate) were performed for demographic characteristics, as well as for knowledge, attitudes, perceptions and practices towards face-mask use. To assess the association between the outcome variables (knowledge on right procedure of wearing face-masks, receipt of information on the use of face-masks, face-mask ownership, face-mask re-use, attitudes, perceptions, and practices) and each explanatory variable, we considered a binary logistic regression which provided crude odds ratios and their corresponding 95% confidence intervals (CIs). Variables with p< 0.05 were all added into the multivariate logistic regression to ascertain significant variables for each outcome. The statistical significance levels were two-sided at p<0.05.

## Results

### Social demographics

A total of 644 participants completed the survey questionnaire hence a response rate of 98%. The average age of the participants was 35.1 years ([SD]:11.0, range 14-71), less than half, 259 (40.2%) were within 24-33 years of age, and 340 (52.8%) were male.

Majority of the participants, 248 (38.5%) were Catholic, more than half, 381 (59.2%) worked in food markets, while 330 (51.2%) walked to their places of work (Table 1).

**Table 1:** Social demographics of study participants.

| Variable | Frequency<br>(N = 644) | Percentage<br>(%) |
| --- | --- | --- |
| Age (years) |  |  |
| **14-23** | 71 | 11.0 |
| **24-33** | 259 | 40.2 |
| **34-43** | 167 | 25.9 |
| **44-53** | 104 | 16.2 |
| **54-63** | 34 | 5.3 |
| **64-73** | 9 | 1.4 |
| Sex |  |  |
| **Female** | 304 | 47.2 |
| **Male** | 340 | 52.8 |
| Education level |  |  |
| **Complete secondary** | 127 | 19.7 |
| **Complete primary** | 65 | 10.1 |
| **Incomplete primary** | 57 | 8.9 |
| **Incomplete secondary** | 191 | 29.7 |
| **No formal education** | 26 | 4.0 |
| **Technical /vocational** | 34 | 5.3 |
| **University/ tertiary** | 144 | 22.4 |
| Religion |  |  |
| **Catholic** | 248 | 38.5 |
| **Moslem** | 110 | 17.1 |
| **Seventh Day Adventist (SDA)** | 11 | 1.7 |
| **Pentecostal/born again** | 78 | 12.1 |
| **Anglican** | 194 | 30.1 |
| **\*Other religions** | 3 | 0.5 |
| Commonly used mode of transport |  |  |
| **Boda boda\*\* (Public)** | 10 | 1.6 |
| Cycling | 16 | 2.5 |
| Motor bike (Private) | 35 | 5.4 |
| Taxi | 203 | 31.5 |
| Private car | 50 | 7.8 |
| Walking | 330 | 51.2 |
| Site |  |  |
| Hospital | 81 | 12.6 |
| Police stations | 182 | 28.3 |
| Food markets | 381 | 59.2 |

\*Other religions included; Isamasiya (Jesus is the Messiah), traditionalists and no religion, \*\*Boda boda is a commercial motorcycle

### Knowledge about COVID-19 and use of face-masks

Nearly all, 642 (99.7%) participants reported having heard about COVID-19, while 635 (98.6%) of the participants reported that they had heard and/or seen messages about the disease. The commonly heard and/or seen message reported were handwashing, 230 (36.2%), social distancing, 135 (21.3%) and wearing of face-masks as a protective measure against COVID-19, 136 (21.4%). Majority of the participants, 512 (80.6%) reported having heard and/or seen the messages on local television stations. Other sources of information about COVID-19 reported by the participants included: local radio stations, 408 (64.3%), family and friends, 93 (14.7%), local newspapers,99 (15.6%), social media platforms, 187 (29.5%), and other internet platforms, 37 (5.8%).

Majority, 437 (67.86%) of the participants reported having received information on how to use face-masks. A large proportion, 353(80.8%) of those who had received the information, received it from local television stations. Other reported sources of the information on how to use face-masks included: local leaders or community health workers (CHWs), 114 (26.1%), social media platforms, 36 (8.2%), and local radio stations, 72 (16.5%). Majority, 496 (77.0%) of the participants also reported that they knew the right procedure or steps of wearing face-masks. Regarding face-mask ownership: majority of the participants, 405 (67.8%) reported owning locally-made, non-medical face-masks, mostly made from single-, 143 (35.3%) or -double, 112 (27.7%) layered cotton fabric (mostly “kitenge” a local fabric printed in various colors and designs).

### Factors associated with knowledge on the right procedure of wearing face-masks and receiving information on the use of face-masks

Bivariate analysis showed that age and receipt of information on face-mask use among the participants were the factors associated with knowledge on the right procedure of wearing face-masks. Individuals between 34-43 years of age (OR: 1.87 95%CI: 1.00-3.50) were 1.87 times more likely to know the right procedure of wearing face-masks compared to individuals between 14-23 years of age. Study participants who had received information on the use of face-masks (OR 6.96 95% Cl: 4.66-10.40) were 6.96 times more likely to be know the right procedure of wearing face-masks compared to those who had never received information on the same.

The bivariate analysis also showed that age, sex, education level and the site of work were the factors associated with receiving information on the use of face-masks. Individuals aged 24-33 years (OR: 2.05, 95% CI: 1.20-3.51), were 2.05 times more likely to receive information on the use of face-masks, those aged 34-43 years (OR: 1.92, 95% CI: 1.09-3.40), were 1.92 times more likely to receive information on the use of face-masks, and those aged 44-53 years (OR: 2.14, 95% CI: 1.14-4.02), were 2.14 times more likely to receive information on use of face-masks compared to those aged 14-23 years of age.

Males (OR: 0.62, 95%CI: 0.44-0.86) were 38% less likely to receive information on the use of face-masks compared to the females. Those with no formal education (OR: 0.28, 95%CI: 0.12-0.66) were 72% less likely to have received the information compared to those who had completed secondary education. Participants who worked in the food markets (OR 0.36,95%CI: 0.19-0.66) were 64% less likely to have received the information while those at police stations (OR: 0.51, 95%CI: 0.26-0.98) were 49% less likely to have received the information than those who worked in the hospital.

After adjusting for confounding, only those that had received information on the use of face-masks (AOR: 6.72, 95%CI:4.47-10.08) were 6.72 times more likely to know the correct procedure of wearing face-masks compared to those that did not receive the information. Furthermore, individuals aged 24-33 years (AOR1.9, 95%CI: 1.08-3.35), 44-53 years (AOR2.12, 95%CI: 1.09-4.14), and those aged 54-63 years (AOR3.39, 95%CI: 1.29-8.89), were more likely to have received information on how to use face-masks compared to those aged 14-23 years. Males (AOR:0.58, 95%CI: 0.40-0.83) were less likely to have received information on the use of face-masks. Those with no formal education (AOR: 0.25, 95%CI: 0.09-0.63), were less likely to have received information on the use of face-masks as compared to those who completed secondary education. Lastly, those who worked in food markets (AOR: 0.47, 95%CI: 0.24-0.93) were also less likely to have received the information as compared to those who worked in hospital (Table 2).

**Table 2.** Factors associated with knowing the right procedure and receipt of information on the use of face-masks

| Demographics |  | Know correct procedure of wearing a mask |  |  |  | Received information on face-mask use |  |  |  |
| --- | --- | --- | --- | --- | --- | --- | --- | --- | --- |
|  |  | Unadjusted OR (95%CI) | P-value | Adjusted OR (95%CI) | p-value | Unadjusted OR (95%CI) | P-value | Adjusted OR (95%CI) | P-values |
| Age (years) |  |  |  |  |  |  |  |  |  |
|  | 14-23 |  |  |  |  |  |  |  |  |
|  | 24-33 | 1.78 (1.00-3.17) | 0.052 | 1.36(0.72-2.57) | 0.341 | 2.05(1.20-3.51) | **0.009** | 1.90(1.08-3.35) | **0.025** |
|  | 34-43 | 1.87(1.00-3.50) | **0.048\*** | 1.50(0.76-2.96) | 0.245 | 1.92(1.09-3.40) | **0.025** | 1.68(0.28-1.19) | 0.091 |
|  | 44-53 | 1.44(0.74-2.80) | 0.286 | 1.04(0.50-2.16) | 0.923 | 2.14(1.14-4.02) | **0.018** | 2.12(1.09-4.14) | **0.027** |
|  | 54-63 | 2.77(0.95-8.11) | 0.061 | 1.95(0.62-6.17) | 0.254 | 2.82(1.12-7.08) | **0.027** | 3.39(1.29-8.89) | **0.013** |
|  | 64-73 | 0.39(0.94-1.56) | 0.181 | 0.60(0.13-2.75) | 0.514 | 0.25(0.05-1.28) | 0.096 | 0.28(0.05-1.52) | 0.139 |
| Sex |  |  |  |  |  |  |  |  |  |
|  | Female |  |  |  |  |  |  |  |  |
|  | Male | 0.730(0.5-1.05) | 0.097 |  |  | 0.62(0.44-0.86) | **0.005** | 0.58(0.40-0.83) | **0.003** |
| Education level |  |  |  |  |  |  |  |  |  |
|  | Complete secondary |  |  |  |  |  |  |  |  |
|  | Complete primary | 1.52(0.70-3.26) | 0.286 |  |  | 1.16(0.60-2.24) | 0.665 | 1.12(0.55-2.28) | 0.761 |
|  | Incomplete primary | 1.05(0.50-2.20) | 0.904 |  |  | 0.57(0.30-1.08) | 0.085 | 0.57(0.28-1.19) | 0.133 |
|  | Incomplete secondary | 0.71(0.42-1.18) | 0.189 |  |  | 0.86(0.53-1.39) | 0.536 | 0.83(0.49-1.40) | 0.483 |
|  | No formal education | 0.70(0.28-1.76) | 0.444 |  |  | 0.28(0.12-0.66) | **0.004\*** | **0.25(0.09-0.63)** | **0.003** |
|  | Technical | 1.19(0.47-3.01) | 0.709 |  |  | 0.93(0.41-2.09) | 0.854 | 0.93(0.41-2.14) | 0.873 |
|  | /vocation<br>al |  |  |  |  |  |  |  |  |
|  | University/<br>tertiary | 1.81(0.98-3.36) | 0.059 |  |  | 1.49(0.87-2.56) | 0.148 | 1.21(0.69-2.12) | 0.515 |
| Religion |  |  |  |  |  |  |  |  |  |
|  | Catholic | 0.37(0.05-2.94) | 0.346 |  |  | 0.48(0.10-2.29) | 0.361 |  |  |
|  | Moslem | 0.21(0.03-1.67) | 0.139 |  |  | 0.36(0.07-1.75) | 0.205 |  |  |
|  | SDA |  |  |  |  |  |  |  |  |
|  | Pentecostal/born again | 0.50(0.06-4.25) | 0.526 |  |  | 0.80(0.16-4.04) | 0.785 |  |  |
|  | Protestant | 0.33(0.04-2.66) | 0.298 |  |  | 0.42(0.09-2.00) | 0.278 |  |  |
|  | Other religions | 1 |  |  |  | 0.44(0.03-7.67) | 0.577 |  |  |
| Site category |  |  |  |  |  |  |  |  |  |
|  | Hospital |  |  |  |  |  |  |  |  |
|  | Police station | 0.86(0.44-1.67) | 0.660 |  |  | 0.51(0.26-0.98) | **0.044\*** | 0.56(0.28-1.12) | 0.102 |
|  | Market | 0.68(0.37-1.26) | 0.220 |  |  | 0.36(0.19-0.66) | **0.001\*** | 0.47(0.24-0.93) | **0.029** |
| Received information on face-mask use |  |  |  |  |  |  |  |  |  |
|  | No |  |  |  |  |  |  |  |  |
|  | Yes | 6.96(4.66-10.40) | **0.000\*** | **6.72(4.47-10.08)** | **<0.001** |  |  |  |  |

### Attitudes and perceptions on COVID-19 and use of face-masks

Majority, 531 (82.5%) of the participants reported that they feared, 267 (41.5%) and were very fearful, 264 (41.0%) of COVID-19. Likewise, majority, 590 (91.6%) of the participants reported that they agreed, 336 (52.2%) and strongly agreed, 297 (46.1%) that acquiring COVID-19 is serious.

Furthermore, majority, 562 (87.3%) of the participants, agreed, 336 (52.2%) and strongly agreed, 226 (35.1%) that face-masks are a good protective measure against COVID-19. With regards to whether or not participants would indefinitely wear face-masks if the COVID-19 threat persisted, majority, 442 (68.6%) reported that they would. Others, 202 (31.4%) reported that they would not, as the majority, 165 (81.7%) thought it is would be an inconvenience. Majority, 568 (88.2%) of the participants also reported that they would easily wear face-masks if everyone in their communities was wearing one.

A large proportion, 531 (82.4%) of the participants reported that they would easily wear face-masks if there were banners and posters available to remind them do so. Others, 432 (81.6%) thought that, the other ways that could remind them about wearing face-masks would be local television and radio stations. More than half, 458 (71.1%) of the participants thought the government’s response to COVID-19 was adequate.

### Factors associated with attitudes and perceptions towards COVID-19 and face-mask use

Bivariate analysis showed that only receiving information on face-mask use was associated to whether one would be comfortable wearing a face-mask indefinitely if COVID-19 persisted. Those who received information on the use of face-masks (OR: 1.58 95% CI: 1.11-2.23) were 1.58 times more likely to be comfortable wearing them indefinitely if COVID-19 persisted.

The bivariate analysis also showed that age, sex, education level and receipt of information on face-mask use were the factors associated to people’s perception on whether a mask is a good protective measure against COVID-19. Participants aged 64 years of age and above (OR:0.18, 95%CI: 0.04-0.80) were 89% less likely to perceive the use of face-masks as a good protective measure against COVID-19 compared to those below 64 years of age. Male participants (OR: 0.61, 95%CI: 0.38-0.97) were 39% less likely to perceive the use of face-masks as a good protective measure against COVID-19 compared to the females. Those who completed primary (OR:3.64, 95%CI:1.03-12.78) were 3 times more likely to perceive the use of face-masks as a good protective measure against COVID-19 compared to those who completed secondary school.

After adjusting for confounders, those aged 64 years and above (AOR:0.17, 95%CI: 0.03-0.82), 83% were less likely to perceive the use of face-masks as good protective measures against COVID-19 compared to those below 64 years of age. Those who received the information on the use of face-masks (AOR:1.83, 95%CI:l.ll-3.02), were more likely to perceive the use of face-masks as good protective measures against COVID-19 compared to those who had never received the same information (Table 3).

**Table 3.**
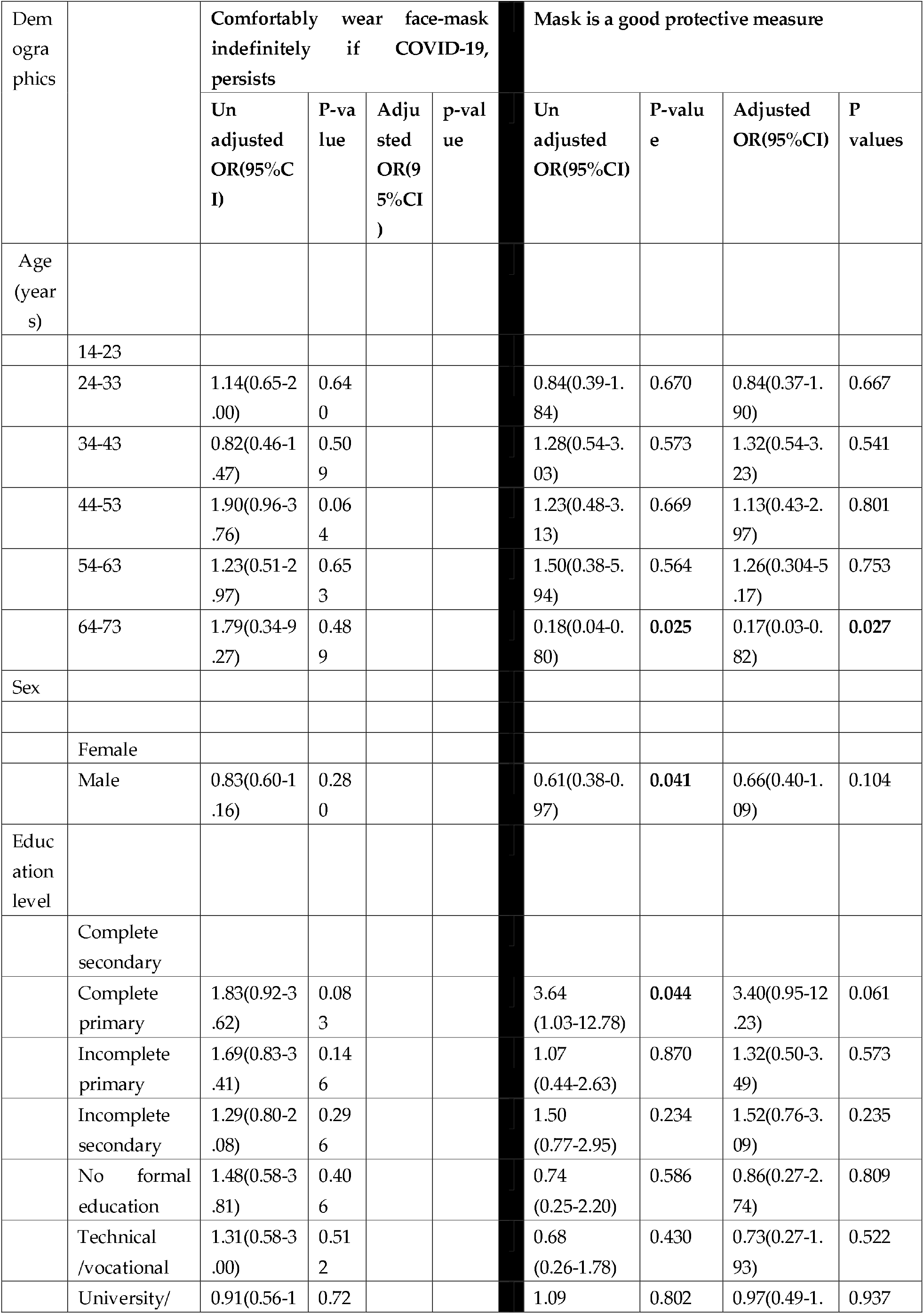
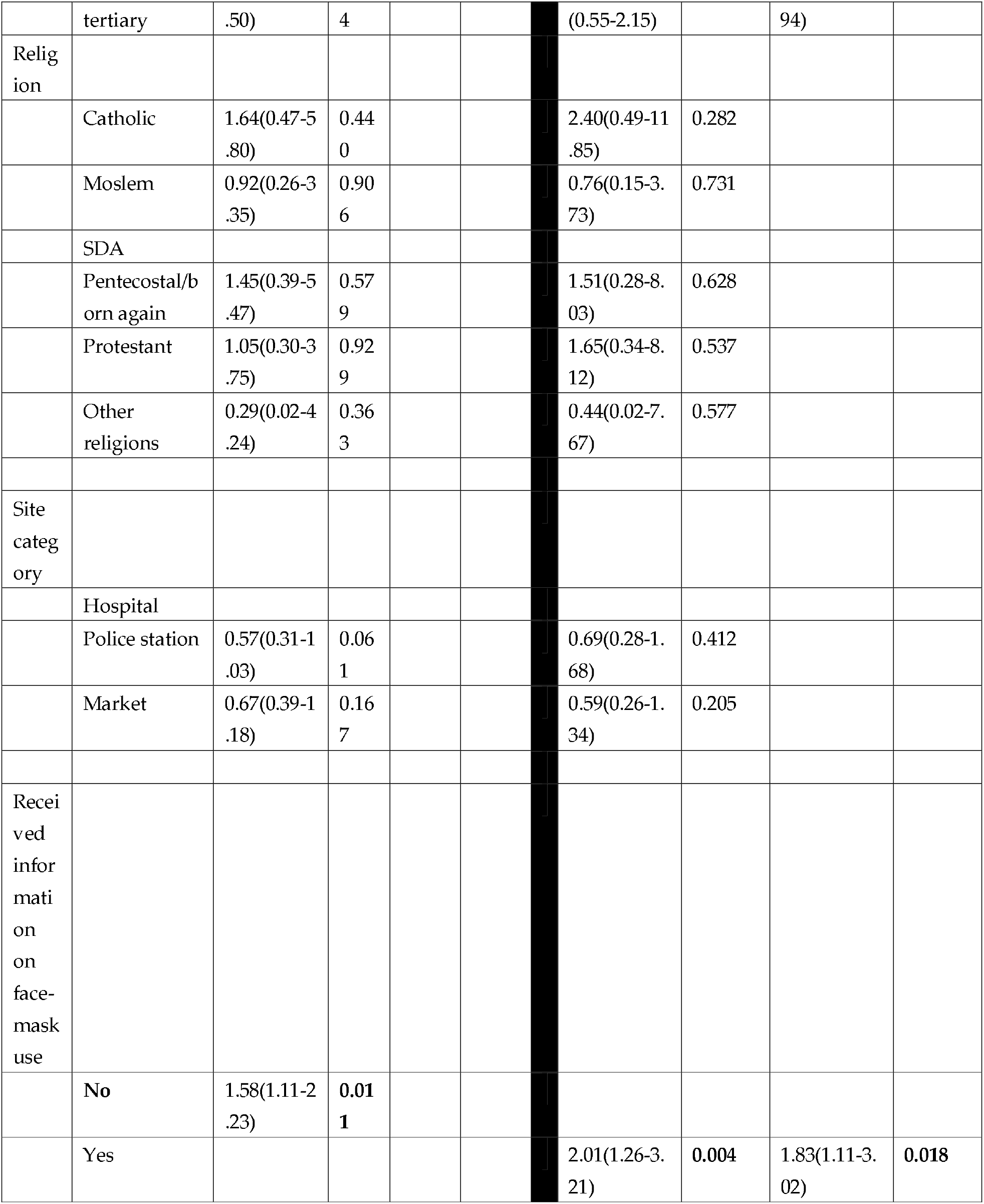
Factors associated to attitudes and perceptions towards COVID-19 and face-mask use11 of 26

### Practices towards the use of face-masks

Almost all, 601 (93.32%) of the participants had done something to protect themselves and their families from COVID-19. Majority, 489 (81.36%) had practiced hand washing with soap and water for at least 20 seconds while more than half, 424 (70.55%) had worn or used face-masks. Majority, 490 (82.1%) of the participants reported that they had re-used their face-masks whether or not they were re-usable. In addition, majority, 203 (41.4%) of those who re-used their face-masks reported that they had done so for one week or less, while a significant number, 113 (23.1%) of participants reported that they had re-used their face-masks for more than one month.

### Factors affecting the practices on the use of face-masks

Bivariate analysis showed that age, site of work and receipt of information on the use of face-masks were the factors associated to ownership of face-masks while education status and site of work were the factors associated to re-use of face-masks. Participants aged 24-33 years (OR:2.78, 95%CI:1.23-6.31), and those within 34-43 (OR:2.60, 95%CI:1.07-6.31) were more likely to own face-masks compared to those aged 14-23 years. Study participants who worked in the food markets (OR:0.34, 95%CI: 0.15-0.78) were 66% less likely to own face-masks compared to those who worked in the hospital. Those who had received information on the use of face-masks (OR:3.44, 95%CI:1.87-6.32) were 3.44 times more likely to own face-masks than those who never received information on the same.

Participants who had completed primary school (OR: 5.36, 95%CI:1.55-18.49), were 5.36 times more likely to re-use their masks compared to those that had completed secondary school and those who had not completed primary school (OR: 3.30, 95%CI: 1.09-10.00) were 3 times more likely to re-use their face-masks compared to those who had completed secondary. Participants that worked in food markets (OR: 4.61 95% CI: 2.47-8.59) were 4.61 times more likely to re-use their face masks compared to those that worked in the hospital.

At multivariate analysis, participants who worked in food markets (AOR: 0.38, 95% CI: 0.16-0.88), were 62% less likely to own face-masks as compared to their counterparts in the hospital. Those who had received the information on the use of face-masks (AOR: 2.85, 95%CI:1.53-5.32), were 2.85 times more likely to own face-masks compared to those who had not received information on the use of face-masks. Furthermore, those who worked in the food markets (AOR: 3.92,95%CI: 1.97-7.82), were 3.92 times more likely to re-use their face-masks as compared to those who worked in the hospital (Table 4).

**Table 4.** Factors affecting the practices on the use of face-masks

| Demographics |  | Own face Mask |  |  |  | Reuse face mask |  |  |  |
| --- | --- | --- | --- | --- | --- | --- | --- | --- | --- |
|  |  | Unadjusted OR(95% CI) | P-value | Adjusted OR(95% CI) | p-value | Unadjusted OR(95% CI) | P-value | Adjusted OR(95% CI) | P-values |
| Age (years) |  |  |  |  |  |  |  |  |  |
|  | 14-23 |  |  |  |  |  |  |  |  |
|  | 24-33 | 2.78(1.23-6.31) | 0.014 | 2.05(0.88-4.76) | 0.097 | 0.74(0.33-1.66) | 0.462 |  |  |
|  | 34-43 | 2.60(1.07-6.31) | 0.035 | 1.61(0.64-4.09) | 0.311 | 0.57(0.24-1.33) | 0.193 |  |  |
|  | 44-53 | 2.54(0.93-6.91) | 0.068 | 1.63(0.58-4.59) | 0.352 | 0.68(0.27-1.67) | 0.394 |  |  |
|  | 54-63 | 6.05(0.75-48.95) | 0.092 | 4.45(0.54-36.92) | 0.167 | 1.12(0.31-4.02) | 0.868 |  |  |
|  | 64-73 | 1.47(0.17-12.92) | 0.730 | 2.21(0.24-20.17) | 0.483 | 0.46(0.08-2.70) | 0.391 |  |  |
| Sex |  |  |  |  |  |  |  |  |  |
|  | Female |  |  |  |  |  |  |  |  |
|  | Male | 0.82(0.45-1.49) | 0.508 |  |  | 1.15(0.76-1.75) | 0.508 |  |  |
| Education level |  |  |  |  |  |  |  |  |  |
|  | Complete secondary |  |  |  |  |  |  |  |  |
|  | Complete primary | 0.49(0.15-1.58) | 0.230 |  |  | 5.36(1.55-18.49) | 0.008 | 2.11(0.57-7.80) | 0.263 |
|  | Incomplete primary | 0.35(0.11-1.11) | 0.074 |  |  | 3.30(1.09-10.00) | 0.034 | 1.14(0.35-3.78) | 0.826 |
|  | Incomplete secondary | 0.49(0.17-1.16) | 0.097 |  |  | 1.48(0.82-2.66) | 0.194 | 0.94(0.49-1.81) | 0.861 |
|  | No formal education | 0.60(0.11-3.13) | 0.540 |  |  | 2.01(0.56-7.25) | 0.286 | 0.73(0.19-2.90) | 0.659 |
|  | Technical /vocational | 1.64(0.19-14.07) | 0.654 |  |  | 1.29(0.48-3.45) | 0.609 | 1.04(0.37-2.94) | 0.932 |
|  | University/ tertiary | 1.14(0.36-3.63) | 0.824 |  |  | 0.81(0.46-1.44) | 0.480 | 0.92(0.49-1.72) | 0.802 |
| Religion |  |  |  |  |  |  |  |  |  |
|  | Catholic | 1.49(0.73-3.07) | 0.277 |  |  | 1.28(0.78-2.09) | 0.325 |  |  |
|  | Moslem | 0.86(0.39-1.92) | 0.720 |  |  | 1.81(0.91-3.59) | 0.090 |  |  |
|  | SDA |  |  |  |  | 1 |  |  |  |
|  | Pentecostal/born again | 1.78(0.58-5.46) | 0.316 |  |  | 1.10(0.55-2.10) | 0.832 |  |  |
|  | Protestant | 1 |  |  |  | 1 |  |  |  |
|  | Other religions | 1 |  |  |  | 1 |  |  |  |
| Site category |  |  |  |  |  | 1 |  |  |  |
|  | Hospital |  |  |  |  | 0.82(0.46-1.46) | 0.503 | 0.78 | 0.424 |
|  | Police station | 1 |  |  |  | 4.61(2.47-8.59) | <0.001 | 3.92(1.97-7.82) | <0.001 |
|  | Market | 0.34(0.15-0.78) | 0.010 | 0.38(0.16-0.88) | 0.025 |  |  |  |  |
| Received information on face-mask use |  |  |  |  |  |  |  |  |  |
|  | No | 1 |  |  |  |  |  |  |  |
|  | Yes | 3.44(1.87-6.32) | <0.001 | 2.85(1.53-5.32) | 0.001 |  |  |  |  |

## Discussion

To the best of our knowledge, this is the first study assessing the knowledge, attitudes and practices (KAP) towards COVID-19 and the use of face-masks among Ugandan high-risk groups. In this study, we analyzed knowledge on the right procedure of face-mask use, receipt of information on the use of face-masks, and face-mask ownership as well as their associated factors. These findings could be useful for public health policy-makers, health workers, and other stakeholders to improve the uptake of face-masks as a key intervention in the prevention of COVID-19 for example through health education among key populations.

In this study, majority of the participants reported having heard about COVID-19, an indication that they were knowledgeable about the disease. The majority of the participants held non-optimistic attitudes and perceptions towards COVID-19. Indeed, many participants reported that they were fearful about the disease, and also agreed that contracting the virus was serious. In light of this, the practices of the participants were very cautious as nearly all reported having done something to protect themselves and their families from COVID-19. These participants reported having practiced hand washing with soap and water for the recommended 20 seconds and having worn face-masks. The participants however believed that the government’s response to COVID-19 had been adequate. This could be attributed to the actions the government had undertaken in the early stages of the pandemic that included: (i) suspension of all public gatherings, (ii) closure of all schools, (iii) suspension of public transport among others (38-40). These actions could have positively affected the perceptions and practices towards COVID-19.

Unlike the finding of a similar study (41), the finding in this study where majority of the participants had reported being knowledgeable about COVID-19 was expected. This is because this study was conducted during the time Uganda’s COVID-19 infections had entered stage three as had been declared by her Ministry of Health in a press release in early June 2020 (42). However, the finding could also be attributed to the efforts that had been pursued by the Ugandan government specifically her Ministry of Health to educate the population about the disease, across several fora such as local television and radio stations. This finding is also similar to that in studies elsewhere that have reported high levels of COVID-19 knowledge in the general population and health-care workers (43-46). Improved knowledge on infectious diseases has been shown to avert negative attitudes while promoting positive preventive practices (46). We also believe that the above finding could be due to the participants’ attitudes and perceptions towards COVID-19. Indeed, majority of the participants reported that they feared COVID-19. Due to the threat of the pandemic and the overwhelming news reports on this public health emergency, these populations could have heard of COVID-19 from various channels of information. These sources of information include: local newspapers, television and radio stations, social media and other internet platforms notably: the official websites and social media accounts of the Uganda’s Ministry of Health and Makerere University, Uganda’s largest and oldest institution of higher learning (47,48).

Uganda has in the past experienced several viral diseases outbreaks such as Ebola during which it has learnt invaluable lessons on how best to deal with these diseases. Indeed, the majority of the population have developed belief in their government’s ability to respond to these diseases, as these responses have been refined overtime (20,49-51). In the case of this study, the belief that the Ugandan government’s response was adequate could be related to the: (1) manner in which the country handled previous viral diseases outbreaks, hence belief already instilled in the Ugandan population but also the unprecedented COVID-19 control measures such as the lockdown of the entire country, (2) willingness to heed to the call sent across by the Ugandan government for concerted efforts from across the country particularly the business community, religious and cultural institutions to: comply with the directives provided by the Ugandan Ministry of Health and cease conducting business, indefinitely suspend religious and cultural gatherings while encouraging their followers to observe all the guidelines provided to prevent the transmission of COVID-19 (52), could have also increased the confidence of the Ugandans, as it demonstrated the belief that the different stakeholders had in the government’s capability to handle the situation, and (3) high knowledge levels about COVID-19 among the target groups could also explain this phenomenon, as increase in knowledge could have been attributed mostly to the efforts of the Ugandan government. Our study established that participants held non-optimistic attitudes and perceptions towards COVID-19 as the majority reported that they were fearful about the disease, and they also agreed that contracting the virus was serious. In light of this, the practices of the participants were very cautious: almost all reported having done something to protect themselves and their families from COVID-19. The practices included having practiced hand washing with soap and water for the recommended 20 seconds, and having acquired and worn face-masks. These strict preventive measures could primarily be attributed their fear of COVID-19, but also, to the strict prevention and control measures that had been implemented by the Ugandan government such as banning of all public gatherings among others. Secondly, they could also be the result of the target populations’ high level of knowledge regarding the seriousness of contracting COVID-19.

Fortunately, the present study like other related studies (41,45,46,53,54), showed that despite the use of face-masks not being a norm in the Ugandan society and the shortage of supply of face-masks due to their high demand as reported elsewhere (43,55), majority of the participants owned and had used face-masks as a protective measure against COVID-19. The participants also reported that they had received information on the use of face-masks via various channels: local leaders and CHWs, local television and radio stations, as well as social media and other internet platforms, and believed that they knew the right procedures of how to use face-masks. This finding is consistent with those of other studies that have showed that: televisions, radios, social media and other internet platforms constitute the major sources of information about COVID-19 (45,46,56). In addition, the transition from television and radios to social media and other internet platforms continues at an unprecedented rate in Uganda. Indeed, the use of smart phones continues to increase across the country, internet connectivity is currently progressing from a luxury for the rich to a felt need for the middle class, and internet cafes are still flourishing throughout the capital city Kampala with lower prices. These developments in the country could explain the increasing use of social media and other internet platforms as sources of information on COVID-19 for the population (57). Also, over time local leaders and CHWs have continued to play a critical role in information dissemination particularly during disease outbreaks in Uganda (58). Similar to a recommendation of another study in the same setting (46), this finding underscores the need to frequently use such media to disseminate COVID-19 related information. In addition, this study underscores the need to utilize local leaders and CHWs in the dissemination of COVID-19 related information, in addition to the different media platforms.

In this study, knowledge about the right procedure of wearing face-masks was related to receipt of information towards the use of face-masks which was related to age, sex, education levels and site of work. The decreased likelihood of receiving information on the use of face-masks was related to the young (24-33 years of age), males, having no formal education, and working in food markets.

As suggested by the findings from previous studies regarding age and gender patterns of risk-taking behaviors (59-61), men and late adolescents (the young) are more likely to engage in risk-taking behaviors. In line with the previous findings, we found a less likelihood of the males and the young receiving information on face-masks use. Males were also less likely to perceive the use of face-masks as a protective measure against COVID-19, hence potentially dangerous practices towards COVID-19.

Our study showed a high level of COVID-19 awareness as well as a high level of knowledge about the right procedure of wearing face-masks among the participants. This finding signifies a positive predictor in curtailing the COVID-19 pandemic within high-risk groups in Uganda. Strictly speaking, our study findings can only be generalized to Ugandan populations of a relatively high socioeconomic status. Considering that educational attainment and occupation are often used as proxy measures of socioeconomic status (41), these findings excluded the underprivileged (i.e. those with no formal education working in the food markets). The likely diminished understanding of the English language and the reduced likelihood of owning either a television set, radio or mobile phone or even accessing the internet and online information resources in these particular populations underscores the need to: (1) pursue research on KAP towards COVID-19 in these populations in Uganda, (2) identify other platforms/means of disseminating knowledge with regards to COVID-19 and practices thereof. Efforts to utilize local leaders and CHWs as well as the dissemination of knowledge pertaining COVID-19 in various local languages could also be pursued.

Unlike the findings of related studies where ownership and use of face-masks was less common (43,46), majority of this study’s participants owned and used face-masks, and believed that the use of the face-masks would protect them from contracting COVID-19. However, this study’s findings are similar to those of other studies (45,62,63). Age and receipt of information on the use of face-masks were the factors that were associated with people’s attitudes and perceptions on whether face-masks were a good protective measure against COVID-19. Participants also reported that they would wear face-masks indefinitely incase the COVID-19 threat persisted, and suggested that with constant reminders (especially via banners and posters, television and radio reminders) and watching others in their settings/communities wearing them, they would continue wearing their face-masks. This finding is consistent with the perspective that face-masks are beyond simply pieces of fabric but rather symbols that serve as constant reminders, and that indicate the presence of a threat(s) (31). This finding suggests that face-masks could be leveraged as symbols that could gradually impact attitudes, perceptions and practices towards COVID-19 in these populations while offering protection against acquiring the virus.

Despite the low certainty evidence as alluded in a number of studies and perspectives (31,64), regarding the protection offered by face-masks in the prevention of COVID-19, our findings on ownership and the use of face-masks by the participants were expected and could be explained by their fear of COVID-19 and the perceived seriousness of contracting the virus. This explanation has also been expounded in the perspective (31), in which expanded masking protocols’ greatest contribution was noted as their role in reducing the transmission of nervousness, over and above whatever role they could play in reducing COVID-19 transmission. The findings on the perceived role of face-masks in preventing the spread of COVID-19 underscore the need to pursue quality, cost-effective research including randomized trials in multiple settings to examine research gaps related to aerosol generating procedures and airborne transmission of SARS-CoV-2, as face-mask use appears to be an acceptable prevention measure to many.

Furthermore, in the absence of research affirming that face-masks do not offer protection against COVID-19. This study’s findings underscore the need for all countries to critically consider the opinions of available studies that have evaluated pre- and asymptomatic-transmission of SARS-CoV-2 and a growing compilation of observational evidence on the use of face-masks by the general public conducted in several countries during the COVID-19 pandemic. In so doing, these countries should adopt the current guidelines provided by WHO and CDC with regards to the use of face-masks in health-care and community settings to: (1) prevent the infected wearer transmitting the infection to others, (2) offer protection to the health wearer against infection, (3) abate circumstances where there could be high risk of exposure to SARS CoV-2 due to intensity of transmission and epidemiology in the population coupled with limited or no capacity to implement other containment measures for example contact tracing, testing and isolation, and care of suspected and confirmed cases; also depending on occupation: individuals working in close contact with the public, (4) offer protection in settings with high population density and settings where individuals are unable to keep a physical distance; particularly those where the risks are greater to ensure a comprehensive approach towards preventing the transmission of COVID-19 (22).

The finding where older participants believed that face-masks were not a good protective measure against COVID-19 may be attributed to their inadequate knowledge about COVID-19, specifically the use of the face-masks as a preventive measure against the disease. This is consistent with another study (65), that reported greater difficulties in accessing novel information, higher likelihood of encountering financial or resource barriers to implement preventive measures among old people, as well as poor neighborhoods and communities.

Unfortunately, some of the participants in this study reported that they could not wear face-masks indefinitely if the COVID-19 threat was to persist, as they found them an inconvenience. This finding could be explained by this study’s other findings in which the receipt of information on the use of face-masks was related to comfort of wearing the face-masks for as long as it was believed necessary; as those who had received the information on the use of face-masks were more likely to be comfortable wearing the face-masks for as long as it was believed necessary. Improved knowledge is critical in shaping people’s behaviour and practices particularly during disease outbreaks because knowledge is partly linked with panic emotion among most populations, which in turn influences their attitudes, perceptions and practices as has been reported in the case of COVID-19 (66). However, improved knowledge in the same populations may not be sufficient to cause behavioral change regarding the use face-masks for extended durations of time. This study’s findings therefore underscore the need to bridge the gap between knowledge and practice by utilizing more interactive and participatory training models developed in a participatory manner involving the different stakeholders for example through focus group discussions as well as field simulations.

In addition, efforts to train high risk populations on the use of face-masks should be encouraged as this would ensure increased ease of using the face-masks as a protective measure against COVID-19. Also, education on other COVID-19 control measures could be disseminated as best alternatives to the adult groups who may have difficulties accepting the use of face-masks as a protective measure against COVID-19.

Regarding the finding where the majority of the participants had re-used their face-masks: majority having had re-used them for up to a week and others for more than one month. Re-use was found associated with education status (having no formal education) and site of work (working in food markets) and this could be explained by: (1) the unavailability or shortage of the face-masks and (2) high costs of the available face-masks in Uganda (67,68). Previous studies have reported the prolonged use and re-use of medical face-masks despite the recommendation for their single use due to their unavailability or shortage especially during pandemics or extended outbreaks and other high demanding situations (69-71). However, the prolonged use or re-use of medical face-masks has also been documented as high-risk practices that could lead to self-contamination of the wearer and hence sources of infection (72).

The limited supply of face-masks and the enforcement of the mandatory wearing of face-masks in all public places by the Ugandan government led to an unprecedented increase in local production of non-medical face-masks. These masks are mostly made up of locally available materials, at both small and large scale as was reported via several local tabloids (73). The locally manufactured face-masks, were mostly single- or - double layered, and had been made out of mostly cotton fabric commonly known as “kitenge”, were cheaper and readily available to the masses. The availability of the cheap locally-made face-mask could explain the finding where the majority of the participants owned and used non-medical face-masks. However, similar to medical masks, the prolonged use or re-use of non-medical face-masks could be high-risk practices that could lead to self-contamination of the wearer and hence sources of infection (72).

This study’s findings underscore the need to sample and perform laboratory testing for both medical and non-medical face-masks commonly circulating on the Ugandan market to assess their safety and fitness-for-use, specific testing could include: microbial filtration efficiency, breathability, splash resistance (synthetic blood), distance dependent fit, microbial cleanliness, and biocompatibility while utilizing available standard operating procedures. This could not only help inform public-health policy makers with regards to the safety and fitness-for-use of the different face-masks circulating on the Ugandan market but could also inform local manufactures on ways to modify their processes so as to locally produce safe medical and non-medical face-masks able to offer protection, while maintaining or promoting health and also a continuous supply of the face-masks. It is worth mentioning that higher COVID-19 knowledge, ownership and use of face-masks and receipt of information on their use scores were found to be significantly associated with a lower likelihood of negative attitudes, perceptions and potentially dangerous practices towards COVID-19 in this study. These findings clearly indicate the importance of improving Ugandans’ COVID-19 knowledge through health education, which may also result in improvements in their attitudes, perceptions and practices towards COVID-19.

Our findings of the demographic factors associated with KAP towards COVID-19 and the use of face-masks are generally consistent with those of previous studies elsewhere on SARS and other viral infectious diseases (41,46,74,75). These findings further suggest that health education interventions would be more effective if they targeted certain demographic groups, particularly, men, the elderly and persons with no formal education.

## Conclusions

Our findings suggest that Ugandan high-risk groups, had good knowledge, optimistic attitudes and perceptions, and relatively appropriate practices towards COVID-19. In addition, good COVID-19 knowledge was associated with optimistic attitudes and appropriate practices towards COVID-19, suggesting that health education programs aimed at improving COVID-19 knowledge are helpful for encouraging optimistic attitudes and perceptions as well as maintaining safe practices especially if they targeted for certain demographic groups, particularly, men, the elderly and persons with no formal education. Furthermore, this study underscores the need to conduct laboratory testing to assess the safety and fitness-for-use for both medical and non-medical face-masks commonly circulating on the Ugandan market.

## Data Availability

The datasets generated and/or analyzed during the current study are not publicly available but are available from the corresponding author on reasonable request.

## Acknowledgments

We thank Ian Amanya, Enock Muzoora, Victo Kyobutungi, Jackie Nabukenya, Simon Peter Otai, Mirembe Annah, Nanfuka Phiona Agatha, Akullo Lilian, Sophie Berna Nansubuga, Stephen Kanyerezi, Grace Kebirungi, Patricia Nsereko, Patricia Nabisubi, Derrick Kitonsa Mugagga, Jeremy Amutuhaire, Aloysious Ssebyala, Richard Ssekitto, Paul Ategyeka, Kester Bataringaya, Muhoozi Michael, Alinda John Vianney, and Jilian Akatwijuka who helped in participant recruitment and data collection, Nagawa Bridget Tamale from Elevate Research Services, who helped in data analysis. We also thank the team at Makerere University Research and Innovations Fund for administrative assistance.

